# Depression with atypical neurovegetative symptoms shares genetic predisposition with immunometabolic traits and alcohol consumption

**DOI:** 10.1101/2020.02.18.20024091

**Authors:** Isabella Badini, Jonathan R.I. Coleman, Saskia P. Hagenaars, Matthew Hotopf, Gerome Breen, Cathryn M. Lewis, Chiara Fabbri

## Abstract

**Background:** Depression is a highly prevalent and heterogenous disorder. This study aims to determine whether depression with atypical features shows different heritability and different degree of overlap with polygenic risk for psychiatric and immuno-metabolic traits than other depression subgroups.

**Methods:** Data included 30,069 European ancestry individuals from the UK Biobank who met criteria for lifetime major depression. Participants reporting both weight gain and hypersomnia were classified as ↑WS depression (N = 1,854) and the others as non-↑WS depression (N = 28,215). Cases with non-↑WS depression were further classified as ↓WS depression (i.e. weight loss and insomnia; N = 10,142). Polygenic risk scores (PRS) for 22 traits were generated using genome-wide summary statistics (Bonferroni corrected p=2.1×10^−4^). Single nucleotide polymorphism (SNP)-based heritability of depression subgroups was estimated.

**Results:** ↑WS depression had a higher polygenic risk for BMI (OR=1.20, [1.15-1.26], p=2.37e-14) and C-reactive protein (OR=1.11, [1.06-1.17], p=8.86e-06) vs. non-↑WS depression and ↑WS depression. Leptin PRS was close to the significance threshold (p=2.99e-04), but the effect disappeared when considering GWAS summary statistics of leptin adjusted for BMI. PRS for daily alcohol use was inversely associated with ↑WS depression (OR=0.88, [0.83-0.93], p=1.04e-05) vs. non-↑WS depression. SNP-based heritability was not significantly different between ↑WS depression and ↓WS depression (14.3% and 12.2%, respectively).

**Conclusions:** ↑WS depression shows evidence of distinct genetic predisposition to immune-metabolic traits and alcohol consumption. These genetic signals suggest that biological targets including immune-cardiometabolic pathways may be relevant to therapies in individuals with ↑WS depression.

## Introduction

Depressive disorders are highly prevalent and a leading cause of global disability and are associated with premature mortality (James et al., 2018) (Kessler & Bromet, 2013). Twin-based heritability of unipolar depression is estimated to be ~37% (Sullivan et al., 2000), with common single-nucleotide polymorphisms (SNPs) explaining ~9% of variation in depression liability (Wray et al., 2018) (Howard et al., 2019). Efforts to identify genetic variants associated with the disease are hindered by the heterogeneity among depressed cases, who can vary greatly in symptom presentation and severity, clinical course and treatment response (Fried & Nesse, 2015). Clinical heterogeneity may also reflect different underlying biological and causal pathways. Increasing evidence suggests that depressive symptoms and subtypes are differentially associated with genetic risk factors that overlap with other disorders (Beijers et al., 2019) (Milaneschi et al., 2016) (Milaneschi et al., 2017). Investigating the clinical and genetic correlates of more homogenous subtypes may improve the understanding of specific aetiological mechanisms and the development of potential treatment targets. Similarly, identifying whether genetic risk for other disorders overlaps with certain characteristics of depression may help to elucidate biological mechanisms underlying common symptom presentations across multiple disorders.

The atypical subtype of major depression is specified in the DSM-5 by the presence of at least two of the following symptoms: hypersomnia, increased appetite and/or weight gain, leaden paralysis, and interpersonal rejection sensitivity (American Psychiatric Association, 2013). Reversed neurovegetative symptoms (hypersomnia, increased appetite and/or weight gain), in particular, have been found to be highly specific and predictive of clinically defined atypical MDD, as they identify patients with similar sociodemographic and clinical correlates as those reported for classification based on the full DSM-criteria (Benazzi, 2002). Classification of atypical depression based on reversed neurovegetative symptoms alone is more feasible in epidemiological studies, many of which have adopted this criterion instead of the full DSM-5 criteria (Lee et al., 2009) (Matza et al., 2003).

Epidemiological studies have shown differences between atypical and non-atypical depression in sociodemographic factors, clinical features, lifestyle factors and comorbidities. Atypical depression has been associated with earlier age of onset, female gender, more severe and recurrent depressive episodes (Agosti & Stewart, 2001) (Blanco et al., 2012) (Brailean et al., 2020). Findings from UK Biobank (UKB) showed higher rates of smoking, social isolation, loneliness, greater exposure to adverse life events and lower rates of moderate physical activity among atypical cases compared to non-atypical cases (Brailean et al., 2020). Atypical depression has also been associated with higher rates of bipolar disorder and psychiatric comorbidity such as anxiety disorders, binge eating disorder and substance abuse (Agosti & Stewart, 2001) (Blanco et al., 2012) (Brailean et al., 2020) (Lee et al., 2009) (Łojko et al., 2015). Physical health comorbidities more strongly associated with atypical cases include higher body mass index (BMI), inflammation, metabolic syndrome and cardiovascular disease (Brailean et al., 2020) (Lasserre et al., 2014) (Milaneschi et al., 2017). Specifically, evidence suggests stronger links between atypical features of increased appetite and/or weight and immunometabolic dysregulations, such as BMI, C-reactive protein (CRP) and leptin (Milaneschi et al., 2017).

There is increasing evidence to suggest partially distinct genetic profiles among depressive subtypes. SNP-based heritability 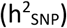 was found to vary across individual depressive symptoms 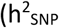 range from 6-9%), and patterns of SNP associations and genetic correlations differed across symptoms (Thorp et al., 2019). A study that classified cases into subtypes according to change in neurovegetative symptoms (i.e. no change, increased or decrease appetite and/or weight [A/W]) found similar SNP-heritability of 10-11% in all groups (Milaneschi et al., 2017). Polygenic risk scoring analyses confirmed that the increased A/W subtype had higher polygenic risk for CRP, BMI, leptin and triglycerides levels than the decreased A/W subtype (Milaneschi et al., 2017) (Milaneschi et al., 2017a) (Milaneschi et al., 2016). Overlapping genetic aetiology between depression and cardiovascular diseases (CAD) was demonstrated (Hagenaars et al., 2019), but no studies investigated if depression with atypical features may have a greater genetic overlap with CAD compared to other depression subtypes.

In the present study, we examined the genetic overlap between depression with atypical features (increased weight and sleepiness: ↑WS) and a range of traits and disorders using polygenic risk scores. Based on previous findings, we hypothesised that ↑WS depression would show similar heritability to depression without ↑WS and higher polygenic risk for immune-cardio-metabolic, substance use and other psychiatric traits compared to depression without ↑WS.

## Materials and methods

### Sample

Individuals who met lifetime criteria for major depression were drawn from the UK Biobank (UKB). UKB is a prospective population-based study of ^~^500,000 individuals recruited across the United Kingdom, aged between 40-69 at baseline (UK Biobank, 2019). A total of 157,387 participants completed an online Mental Health Questionnaire (MHQ) assessing self-reported psychiatric symptoms corresponding to clinical diagnostic criteria and self-reported professional diagnoses (K. A. S. Davis et al., 2020). Genome-wide genetic data has been collected on all UKB participants (Bycroft et al., 2018), as detailed below. All participants provided written informed consent and all procedures contributing to this work comply with the ethical standards of the relevant national and institutional committees on human experimentation and with the Helsinki Declaration of 1975, as revised in 2008.

### Measures

Lifetime psychiatric diagnoses were assessed in the MHQ using the Composite International Diagnostic Interview Short Form (CIDI-SF) (Kessler et al., 1998). Criteria for lifetime major depressive episode were in accordance with DSM-V. The full CIDI is a validated measure of depression, demonstrated to have good concordance with direct clinical assessment (Haro et al., 2006).

Cases reporting both weight gain (data field 20536) and hypersomnia (data field 20534) were classified as ↑WS depression (N = 1,854), and the remaining cases classified as depression without ↑WS (N = 28,215). From this group, depression with both weight loss and decreased sleep (data fields 20533 and 20535) was defined as ↓WS depression (N = 10,142). These definitions used the same coding conventions as those used in a previous study in the UKB (Brailean et al., 2020), however we preferred to avoid the terms atypical and typical depression to avoid confusion with the standard nosological classification. We did not consider variations in appetite to distinguish the subgroups, since the available measure (data field 20511) did not differentiate between hypophagia and hyperphagia.

### Genotyping and quality control

Genetic data came from the full release of the UKB data (N = 488,377; (Bycroft et al., 2018). Genotyping was performed using two highly-overlapping arrays covering ^~^800,000 markers (UK Biobank Axiom Array Content Summary). Autosomal genotype data underwent centralised quality control to adjust for possible array effects, batch effects, plate effects, and departures from Hardy-Weinberg equilibrium (HWE; (Bycroft et al., 2018)). Variants for this analysis were limited to common variants (minor allele frequency > 0.01) that were directly genotyped. SNPs were further excluded based on missingness (> 0.02) and on HWE (p < 10-8). Individuals were removed for high levels of missingness (> 0.05) or abnormal heterozygosity (as defined during centralised quality control), relatedness of up to third-degree kinship (KING r < 0.044 (Manichaikul et al., 2010)), or phenotypic and genotypic gender discordance (phenotypic males with F_x_<0.9, phenotypic females with F_x_>0.6). Population structure within the UK Biobank cohort was assessed using principal component analysis, with European ancestry defined by 4-means clustering on the first two genetic principal components (Warren et al., 2017). Among respondents to the MHQ, 95% were of European ancestry and therefore individuals from other ancestries were excluded from further analyses to maximise statistical power. After quality control, the final sample of respondents to the MHQ consisted of 126,522 individuals with genotype data.

### Statistical analysis

#### Polygenic risk scores

Polygenic risk scores were calculated based on GWAS summary statistics for 22 traits reflecting the hypothesis formulated in the Introduction, including major psychiatric disorders, personality traits, substance use related traits, cardio-metabolic traits and C-reactive protein (CRP) (Supplementary Table 1). There was no overlap between the samples included in these GWASs and the sample included in this study, except a very marginal overlap with the GWAS of anorexia nervosa (349/16,992 (2%) of cases included in (Watson et al., 2019)).

PRS were calculated using PRSice v.2 (Choi & O’Reilly, 2019) (Euesden et al., 2015). PRSice computes scores in an independent (target) sample by calculating the weighted sum of trait-associated alleles using summary data from GWAS discovery samples. SNPs in linkage disequilibrium (r2 ≥ 0.1 [250-kb window]) were removed using the clumping procedure. We used the default average option that calculates the ratio between the PRS and the number of alleles included in each individual and PRS were standardised (mean=0, SD=1). PRS were calculated at 11 p-value thresholds P_T_ (5e-8, 1e-5, 1e-3, 0.01, 0.05, 0.1, 0.2, 0.3, 0.4, 0.5, 1) and the most predictive P_T_ was selected. Logistic regression models were used to estimate associations between ↑WS depression vs. non-↑WS depression and each PRS adjusting for covariates of six genetic ancestry principal components, centre and batch effects. We estimated the proportion of variance explained by PRS on the observed and liability scale (Lee et al., 2012), considering a range of possible values of prevalence among depressed cases (Łojko & Rybakowski, 2017) (Levitan et al., 1997). For PRS of traits associated with ↑WS depression we calculated: 1) the OR of ↑WS depression vs. depression without ↑WS for each decile of the PRS, taking the first decile as reference and 2) if the effect was comparable when considering each of the symptoms separately (↑weight and ↑ sleep vs. no increase in weight or sleep). We estimated differences between PRS results of different comparisons by comparing their estimates (E1 and E2) and SE (SE1 and SE2) using a 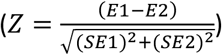. The code used for these analyses is available as supplementary material (code_used_for_analyses).

A Bonferroni correction was applied to account for multiple testing, providing a required significance level of p < 2.1e-4 (the PRS of 17 traits were analysed at 11 P_T_, while for 5 traits there were no SNPs with p < 5e-8 and 10 thresholds were tested [0.05/(11 x 17 + 10 x 5)= 2.1e-4]). The use of Bonferroni correction is conservative, since the different P_T_ are highly correlated.

#### SNP-based heritability

Using genome-wide complex trait analysis software v.1.93.1beta (GCTA) (J. Yang et al., 2011), genetic relationship matrix-restricted maximum likelihood (GREML) methods (Lee et al., 2011) were used to estimate the variance in liability attributable to the additive effects of all SNPs 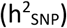 for ↑WS depression, ↓WS depression and depression not falling in these two groups. A random subset of 8,000 healthy controls were selected for the analyses among those who completed the MHQ. This number of controls was selected because it provided adequate power (at least 80%) to estimate heritability considering an expected heritability of 0.10 (Milaneschi et al., 2017), using GCTA-GREML Power Calculator (Visscher et al., 2014). The genetic relationship matrix (GRM) was adjusted for incomplete tagging of causal SNPs and we excluded related individuals using a grm-cutoff of 0.05. We calculated the genetic correlation between depression subgroups using bivariate GREML with independent subsets of 8,000 healthy controls for each depression subgroup.

We also calculated 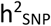 using GCTB (Genome-wide Complex Trait Bayesian analysis) Bayes S method, which estimates polygenicity from the data (i.e., the proportion of SNPs with nonzero effects, Pi). GCTB also calculates the relationship between effect size and MAF (S) which can be used to detect signatures of natural selection (Zeng et al., 2018). We used the standard settings of 21,000 simulations with the first 1,000 as burn-in and standard initial S and Pi values.

GCTA-GREML and GCTB Bayes S analyses were adjusted for the same covariates included in the PRS analysis, but we also adjusted for BMI because there was a significant difference in BMI between cases with ↑WS depression (30.55±5.78), ↑WS depression (25.93±4.41) and healthy controls (26.45±4.14) (p=1.51e-221 and p=1.25e-154, respectively); heritability estimates would be confounded by this variable which was demonstrated to have heritability of about 28% (Zeng et al., 2018).

Possible differences among 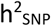 of depression subgroups were compared by a Z-test as explained for PRS results. 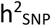 estimates were transformed to a liability scale (S. H. Lee et al., 2012) and we reported heritability considering a range of plausible prevalence values (Lim et al., 2018) (Łojko & Rybakowski, 2017) (Levitan et al., 1997). The code used for these analyses is available as supplementary material (code_used_for_analyses).

#### Sensitivity analyses

Analyses were repeated 1) excluding cases with probable bipolar disorder or missing information for this variable (wider bipolar disorder definition, as described in (Brailean et al., 2020), n=1747) and schizophrenia or missing information for this variable (n=61); 2) comparing ↑WS depression with ↑WS depression (rather than non-↑WS depression); and 3) comparing both ↑WS depression and ↑WS depression with healthy controls who completed the MHQ (n=64,604) (Figure 1).

**Figure 1:**
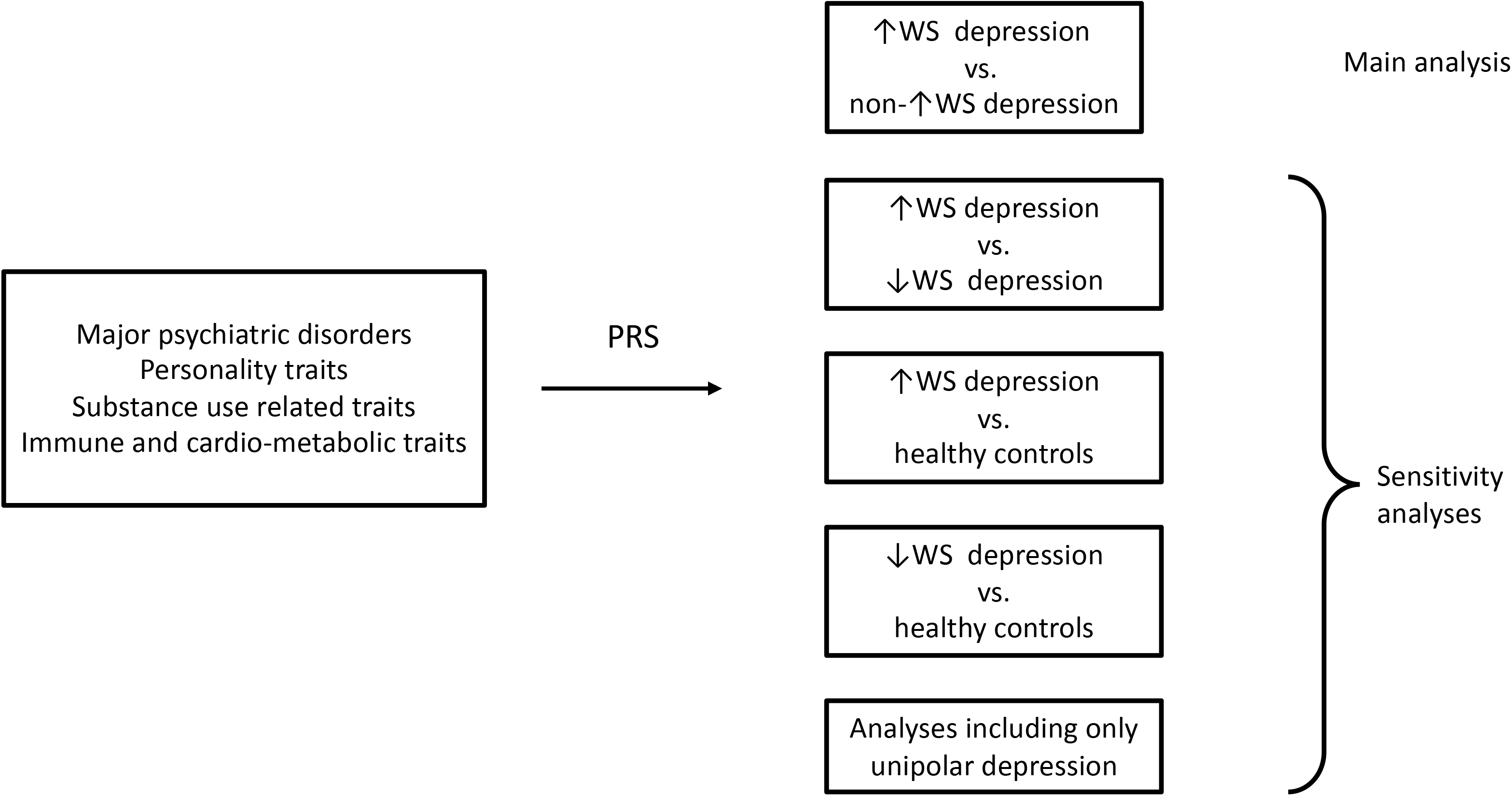
flow chart of the performed analyses. ↑WS = depression with increased weight and sleepiness; ↓WS depression = depression with decreased weight and insomnia.

## Results

### Sample characteristics

The total sample comprised of 30,069 participants who met criteria for lifetime major depressive episode, including 1,854 cases classified as ↑WS depression and 28,215 classified as depression without ↑WS. Non-↑WS cases were further classified as ↓WS depression (N = 10,142). The ↑WS group was 75% female and had a mean age of 59.99 (*SD* = 7.09) years, compared to the non-↑WS group which was 68% female and had a mean age of 62.50 (7.53) years. The ↑WS depression group was 75% female and had a mean age of 62.55 (7.49). Further description of the clinical-demographic features of the subtypes can be found in a previous paper in UKB that used the same classification criteria (Brailean et al., 2020).

### Polygenic risk analysis

Traits where the PRS were significantly associated with ↑WS depression compared to non-↑WS depression included BMI (OR=1.20 [1.15-1.26], p=2.37e-14), CRP (OR=1.11 [1.06-1.17], p=8.86e-06), daily alcohol use (OR=0.88 [0.83-0.93], p=1.04e-05) and MDD (OR=1.10 [1.05-1.15], p=1.14e-04), as shown in Table 1 (see Supplementary Table 2 for all results). The OR for ↑WS depression was 2.03 (1.62-2.55), 1.36 (1.10-1.68), 0.62 (0.49-0.79) and 1.30 (1.04-1.61) in the 10^th^ PRS decile vs. the 1^st^ PRS decile for BMI, CRP, daily alcohol use and MDD, respectively (Figure 2). Results were consistent between different p_T_ (Supplementary Figure 1). The same direction of effect was observed when comparing cases with weight gain vs. cases without weight gain, with similar effect size for alcohol daily use, MDD, leptin and CRP PRS, while larger effect size for BMI PRS (z=2.12, p=0.03). These PRS showed the same direction of effect though not significant when considering depression with hypersomnia compared to depression without this symptom (Supplementary Table 2C). The results were consistent when comparing ↑WS depression with ↓WS depression and ↑WS depression with healthy controls (Supplementary Table 3 and 4). Interestingly, the effect of BMI PRS was very close to the significance threshold for being inversely associated with ↓WS depression vs. healthy controls (p=1.24e-04) while leptin, CRP and alcohol daily use PRS had no effect (Supplementary Table 5).

**Table 1.** Polygenic risk scores associated with depression with atypical features (↑WS depression) compared with depression without atypical features. Results are shown for the p_t_ threshold attaining the lowest p-value.

| Polygenic Risk Score | Reference | Sample size | $P_T$ | $p$ | OR (95% CI) | $R^2$ |
| --- | --- | --- | --- | --- | --- | --- |
| BMI | (Locke et al., 2015) | 322,154 | 0.001 | 2.37e-14* | 1.20 (1.15 – 1.26) | 0.00520 |
| Leptin | (Kilpeläinen et al., 2016) | 31,816 | 0.001 | 2.99e-04 | 1.09 (1.04 – 1.14) | 0.00117 |
| CRP | (Ligthart et al., 2018) | 204,402 | 0.4 | 8.86e-06* | 1.11 (1.06 – 1.17) | 0.00177 |
| Daily alcohol use | (Schumann et al., 2016) | 70,460 | 0.4 | 1.04e-05* | 0.88 (0.83 – 0.93) | 0.00174 |
| Major depressive disorder | (Wray et al., 2018) | 143,265 | 0.3 | 1.14e-04* | 1.10 (1.05-1.15) | 0.00133 |

Note. $R^2$ estimated on the observed scale as the results are referred to case only comparisons.
\*Significant associations ( $p < 2.1e-4$ ).

**Figure 2:**
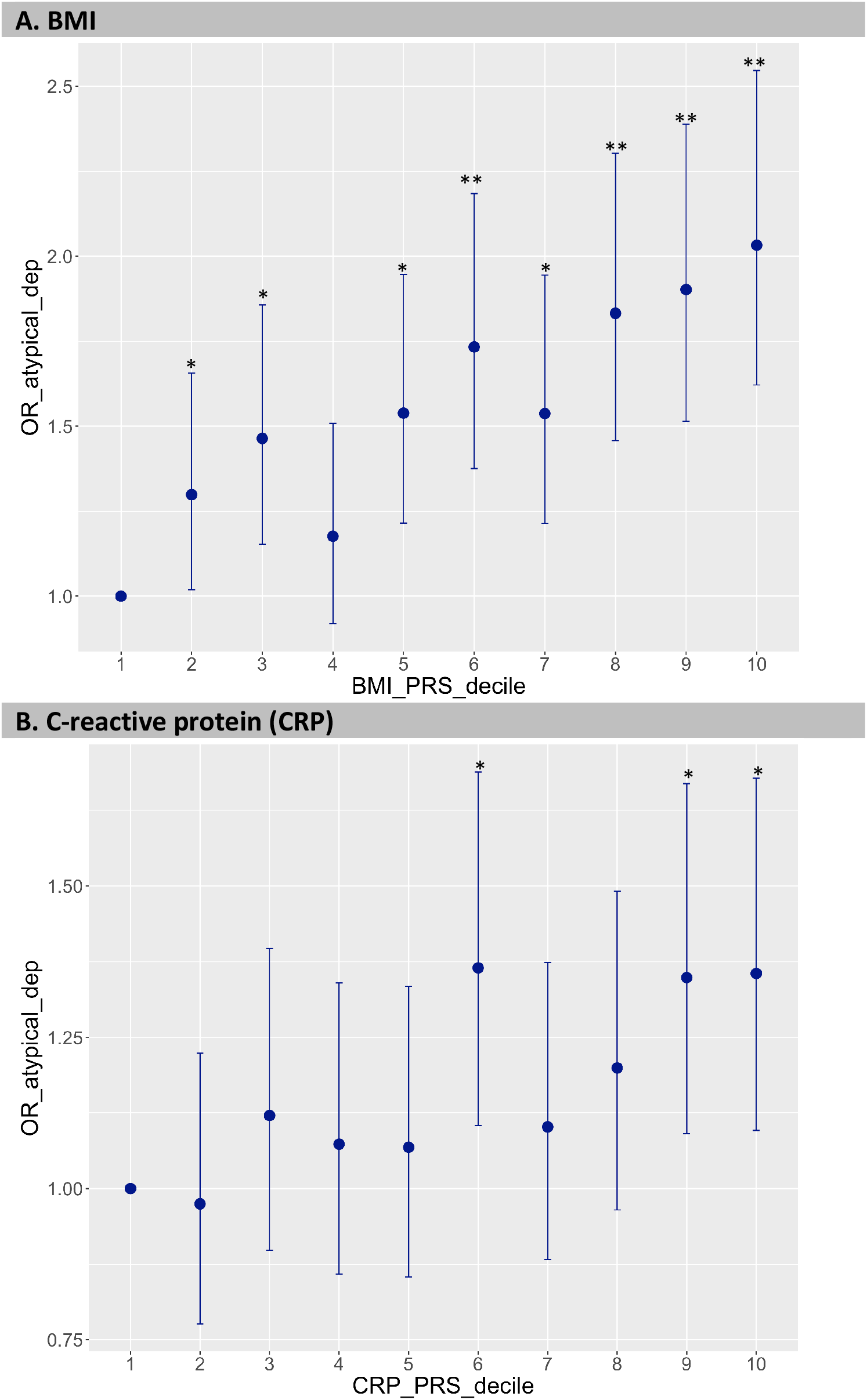
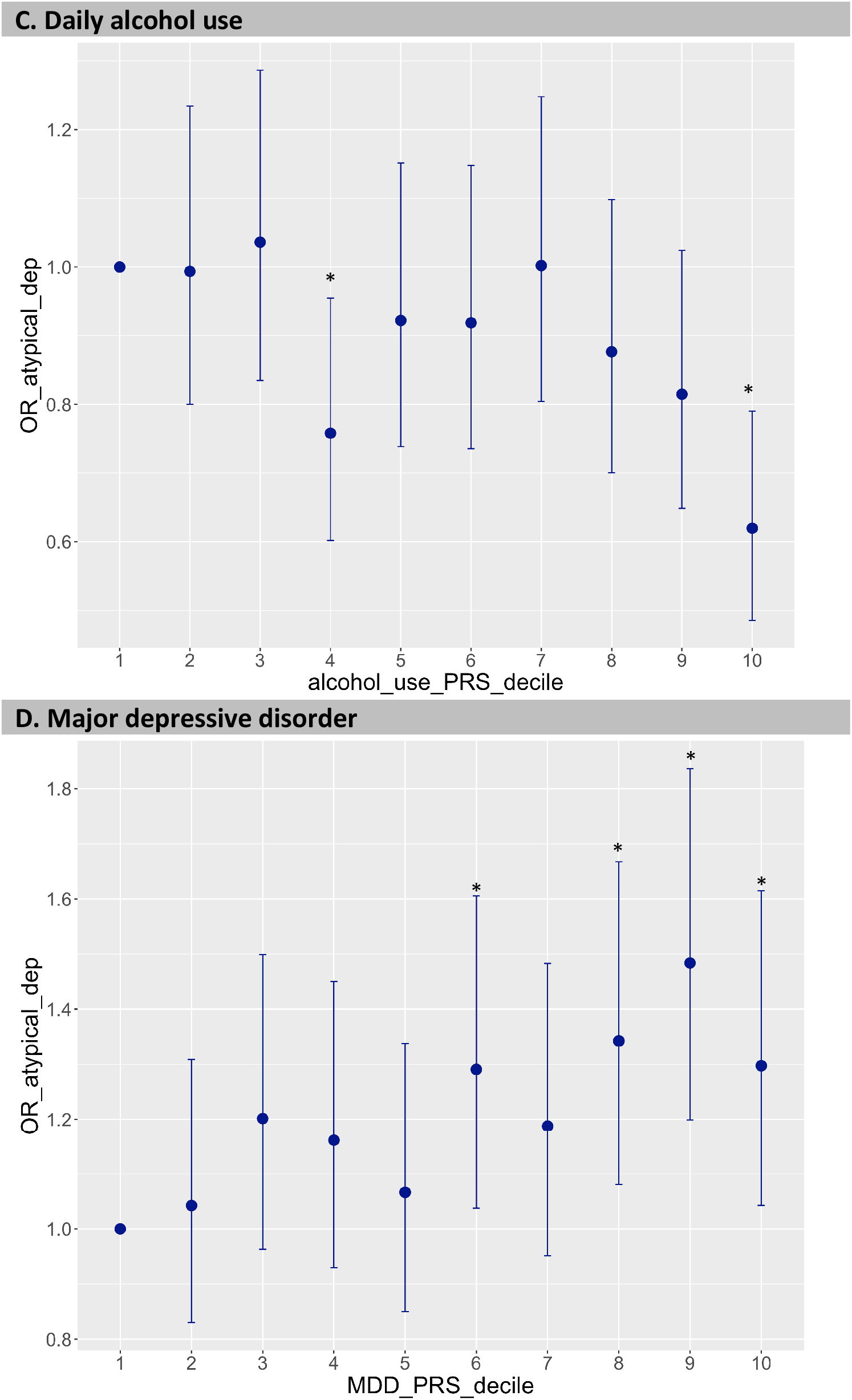
Decile plots showing the odds ratio of ↑WS depression (depression with atypical features) vs. depression without ↑WS for BMI PRS (**A**), C-reactive protein (CRP) PRS (**B**), daily alcohol use PRS (**C**) and major depressive disorder (MDD) PRS (**D**). *p<0.05; **p<1e-05.

The association between ↑WS vs. depression without ↑WS and leptin PRS was close to the significance threshold (OR=1.09 [1.04 - 1.14], p=4.13e-04) but disappeared when considering GWAS summary statistics of leptin adjusted for BMI. Nominal significant associations (p < 0.05) included PRS for type 2 diabetes, coronary artery disease, triglycerides, total HDL cholesterol, and ischemic stroke (Figure 3; Supplementary Table 2). The PRS for coronary artery disease reached statistical significance for association with both ↑WS depression and ↓WS depression when the comparator group was healthy controls, in addition to triglycerides PRS only for ↑WS depression vs. controls. Case-healthy controls analyses showed expected associations with the PRS of psychiatric traits and contributed to clarifying the effect of alcohol use-related traits PRS: higher PRS for alcohol dependence but not daily alcohol use increased the risk of ↓WS depression, while ↑WS depression was still associated with lower daily alcohol use PRS (Supplementary Table 4-5, Figure 4). Sensitivity analysis revealed very similar results when excluding cases with probable bipolar disorder and schizophrenia or missing information for these variables (n=196 and n=12 among patients with ↑WS depression; n=1551 and n=49 among those with non-↑WS depression; Supplementary Figures 2-4).

**Figure 3:**
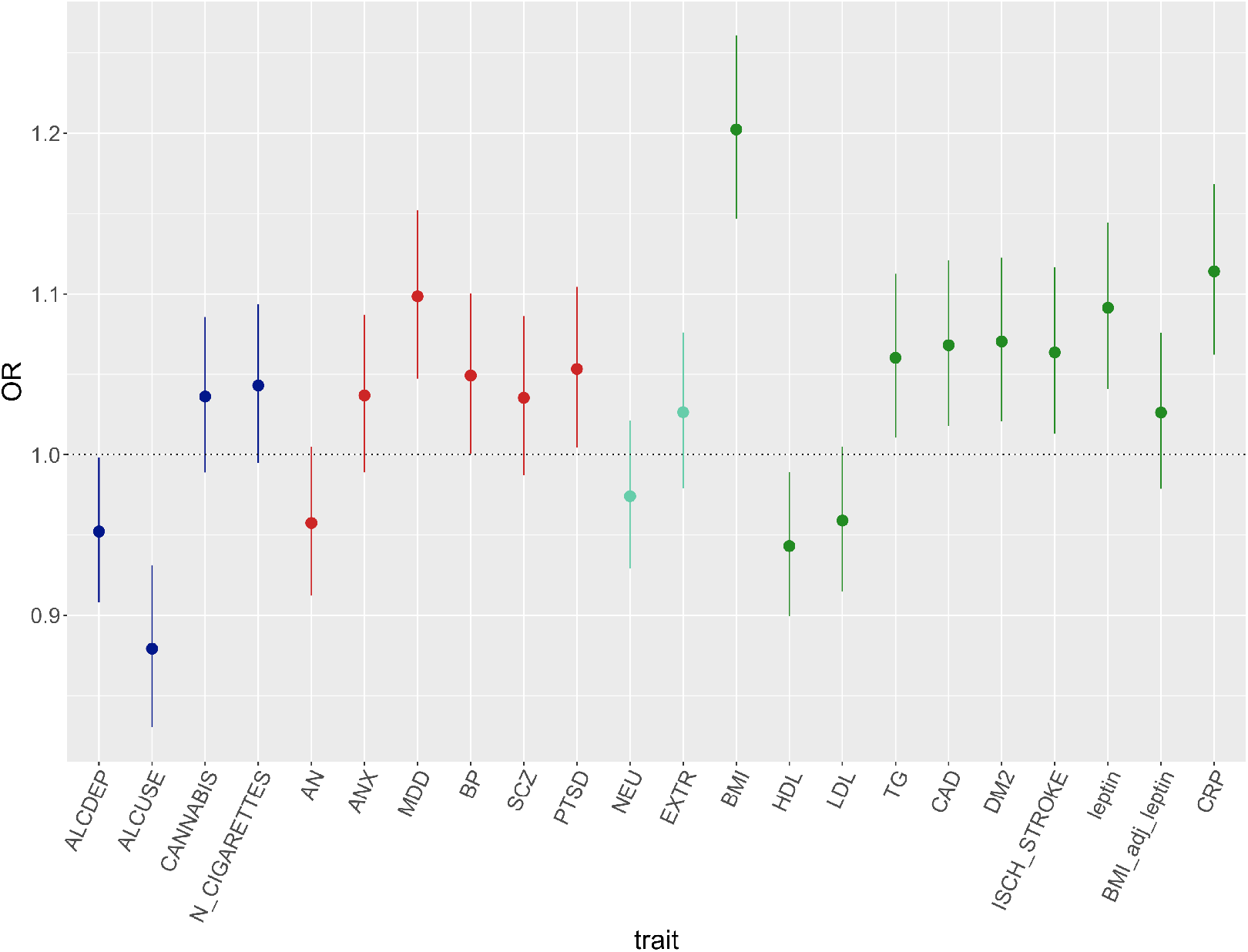
PRS odds ratio (OR) and 95% confidence intervals of ↑WS depression vs. depression without ↑WS. Colors indicate different groups of traits (substance related disorders, major psychiatric disorders, personality traits, immune metabolic traits). BP=bipolar disorder; SCZ=schizophrenia; ANX=anxiety disorders; PTSD=post-traumatic stress disorder; AN=anorexia nervosa; ALCDEP=alcohol dependence; ALCUSE=daily alcohol use; N_CIGARETTES=n cigarettes per day; CANN=cannabis use lifetime; EXTR=extraversion; NEU=neuroticism; DM2=type 2 diabetes mellitus; CAD=coronary artery disease; ISCH_STROKE=ischemic stroke; TG=triglycerides; LDL=total LDL cholesterol; HDL=total HDL cholesterol; BMI=body max index; BMI_adjust_leptin=leptin adjusted for BMI; CRP=C-reactive protein.

**Figure 4:**
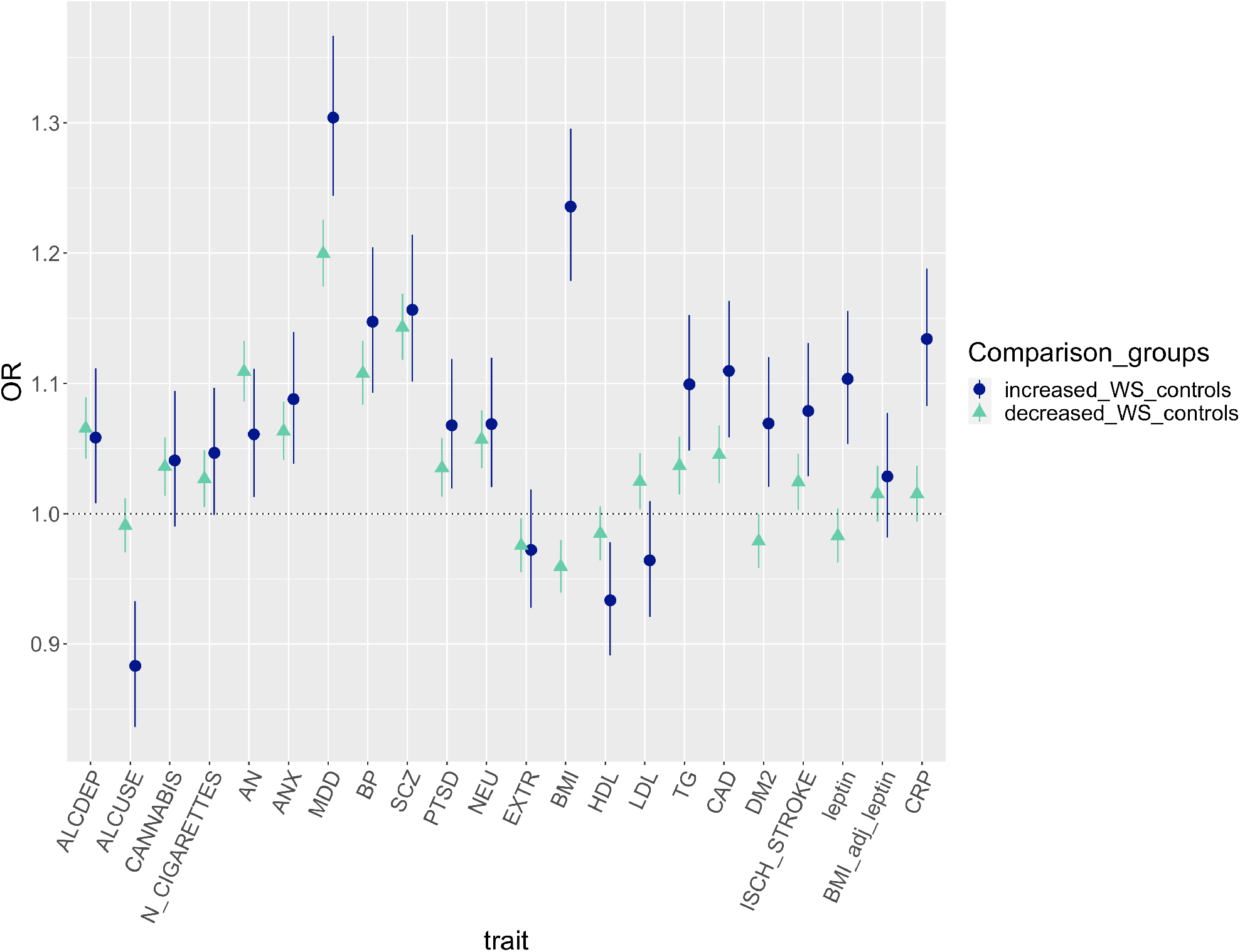
PRS odds ratio (OR) and 95% confidence intervals of ↑WS depression and ↓WS depression compared with healthy controls. BP=bipolar disorder; SCZ=schizophrenia; ANX=anxiety disorders; PTSD=posttraumatic stress disorder; AN=anorexia nervosa; ALCDEP=alcohol dependence; ALCUSE=daily alcohol use; N_CIGARETTES=n cigarettes per day; CANN=cannabis use lifetime; EXTR=extraversion; NEU=neuroticism; DM2=type 2 diabetes mellitus; CAD= coronary artery disease; ISCH_STROKE=ischemic stroke; TG= triglycerides; LDL=total LDL cholesterol; HDL=total HDL cholesterol; BMI=body max index; BMI_adjust_leptin=leptin adjusted for BMI; CRP=C-reactive protein.

### SNP-based heritability

SNP-based heritability 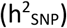 on the liability scale for ↑WS depression was estimated to be 0.14 (95% CI 0.03-0.25) using GCTB-Bayes S and 0.21 (95% CI 0.07-0.35) using GCTA-GREML (Table 2). 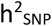 was 0.12 (95% CI 0.08-0.16) and 0.16 (95% CI 0.10-0.22) for ↑WS depression using the two methods, respectively. GCTA-GREML 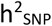 was not significantly different comparing ↑WS depression with ↓WS depression (z=0.57, p=0.56), depression with weight gain (z=1.09, p=0.27), depression with weight loss (z=0.75, p=0.46) or depression without ↑WS or ↓WS (z=1.25, p=0.21). GCTB-Bayes S estimates were not significantly different compared with GCTA-GREM estimates for any depression subgroup (Table 2). The estimated proportion of SNPs contributing to 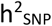 ranged from 2% to 4%; S estimation was negative for all traits but not significantly different from zero, except for depression with weight gain (z=2.36, p=0.018), which however did not survive Bonferroni correction for five tests (Supplementary Figure 5).

Genetic correlations were 0.54 (SE=0.14) for ↑WS depression and ↑WS depression, 1.05 (SE=0.15) for ↑WS depression and depression without ↑WS or ↓WS, and 0.74 (SE=0.14) between ↓WS depression and depression without ↑WS or ↓WS.

**Table 2:**
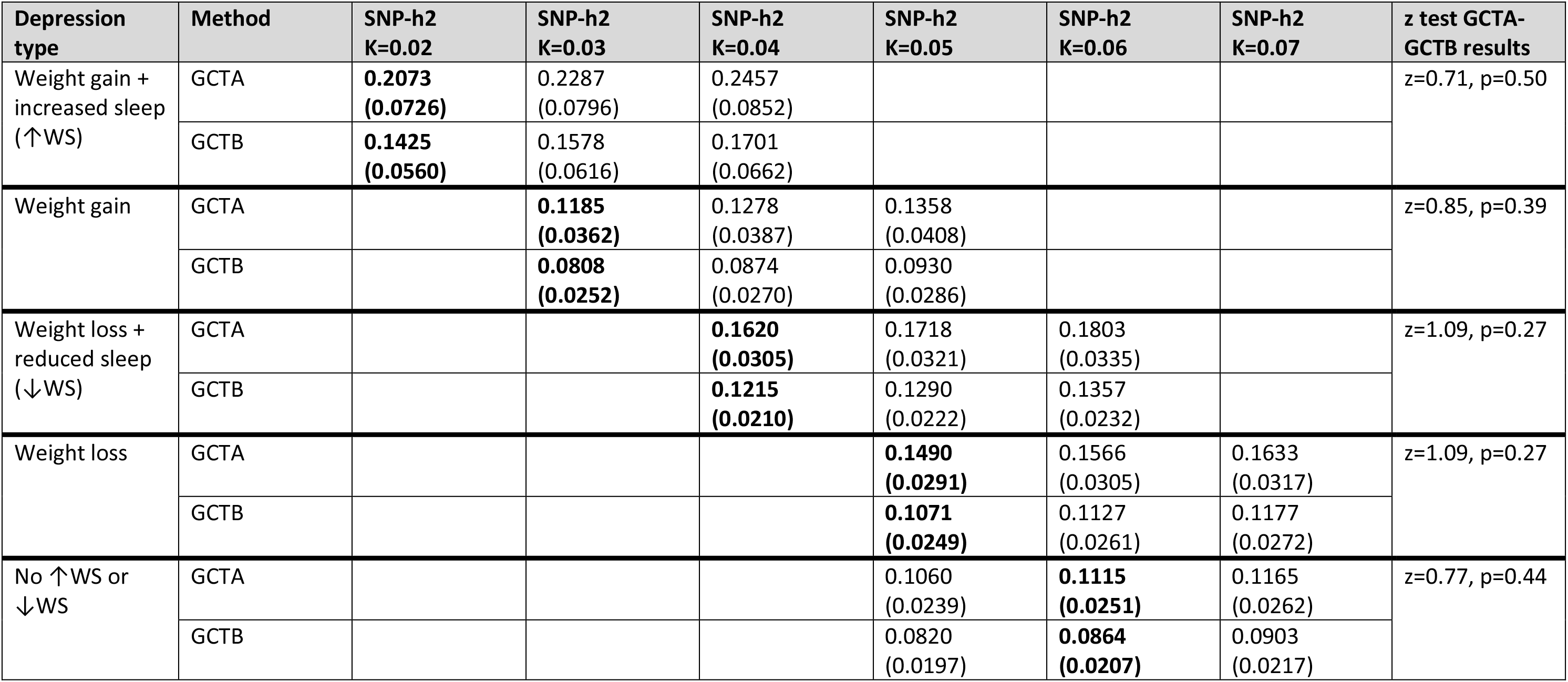
SNP heritability (SNP-h2) of depression with weight gain and hypersomnia (↑WS depression), depression with weight loss and reduced sleep (↓WS depression), major depression not falling in any of these two groups (no ↑WS or ↓WS), depression with weight gain or weight loss. Heritability and SE in parenthesis are reported on the liability scale according to different possible values of prevalence (K) of each subtype in the population. The most credible values of population prevalence are in bold and a Z-test was performed to compare the estimate and SE of GCTA-GREML and GCTB Bayes S results.

| Depression type | Method | SNP-h<sup>2</sup><br>K=0.02 | SNP-h<sup>2</sup><br>K=0.03 | SNP-h<sup>2</sup><br>K=0.04 | SNP-h<sup>2</sup><br>K=0.05 | SNP-h<sup>2</sup><br>K=0.06 | SNP-h<sup>2</sup><br>K=0.07 | z test GCTA-GCTB results |
| --- | --- | --- | --- | --- | --- | --- | --- | --- |
| Weight gain + increased sleep (↑WS) | GCTA | **0.2073**<br>**(0.0726)** | 0.2287<br>(0.0796) | 0.2457<br>(0.0852) |  |  |  | z=0.71, p=0.50 |
|  | GCTB | **0.1425**<br>**(0.0560)** | 0.1578<br>(0.0616) | 0.1701<br>(0.0662) |  |  |  |  |
| Weight gain | GCTA |  | **0.1185**<br>**(0.0362)** | 0.1278<br>(0.0387) | 0.1358<br>(0.0408) |  |  | z=0.85, p=0.39 |
|  | GCTB |  | **0.0808**<br>**(0.0252)** | 0.0874<br>(0.0270) | 0.0930<br>(0.0286) |  |  |  |
| Weight loss + reduced sleep (↓WS) | GCTA |  |  | **0.1620**<br>**(0.0305)** | 0.1718<br>(0.0321) | 0.1803<br>(0.0335) |  | z=1.09, p=0.27 |
|  | GCTB |  |  | **0.1215**<br>**(0.0210)** | 0.1290<br>(0.0222) | 0.1357<br>(0.0232) |  |  |
| Weight loss | GCTA |  |  |  | **0.1490**<br>**(0.0291)** | 0.1566<br>(0.0305) | 0.1633<br>(0.0317) | z=1.09, p=0.27 |
|  | GCTB |  |  |  | **0.1071**<br>**(0.0249)** | 0.1127<br>(0.0261) | 0.1177<br>(0.0272) |  |
| No ↑WS or ↓WS | GCTA |  |  |  | 0.1060<br>(0.0239) | **0.1115**<br>**(0.0251)** | 0.1165<br>(0.0262) | z=0.77, p=0.44 |
|  | GCTB |  |  |  | 0.0820<br>(0.0197) | **0.0864**<br>**(0.0207)** | 0.0903<br>(0.0217) |  |

## Discussion

This study examined the genetic overlap between depression with atypical features (↑WS) and a range of traits and disorders using polygenic risk scores in 30,069 cases with major depression from the UKB. The findings showed that persons with higher BMI PRS, CRP PRS and MDD PRS are more likely to have ↑WS depression vs. non-↑WS depression and ↓WS depression, while those with lower PRS for alcohol daily use were more likely to have non-↑WS depression rather than ↑WS depression. Associations with these PRS were consistent among different p_t_. The effect of BMI PRS and CRP PRS on the risk of ↑WS depression was similar when taking healthy controls as comparator group, while it was in the opposite direction or absent, respectively, when we compared ↓WS depression vs. healthy controls. The analyses of individual symptoms (weight changes and sleep changes) showed that BMI, non-BMI adjusted leptin and CRP PRS had a higher effect size on depression with weight increase than sleep increase, but not the PRS of alcohol daily use and MDD; however, the direction of the effect on weight and sleep increase was the same for all these PRS, suggesting that the symptoms of weight gain and sleepiness in depression have shared rather than divergent genetics. The PRS of coronary artery disease and triglycerides was associated with ↑WS depression vs. healthy controls, in line with nominal associations between the PRS of other cardio-metabolic traits (e.g. type 2 diabetes) and the risk of ↑WS depression compared to other depression subgroups as well as healthy controls. The PRS for alcohol dependence was associated with the risk of ↑WS depression compared to healthy controls but not for ↑WS depression compared to healthy controls, though the direction of the effect was the same. Interestingly, the PRS of alcohol daily use was similar between ↑WS depression and healthy controls, but significantly lower in ↑WS depression, suggesting that the pathogenesis of alcohol use disorders may involve different mechanisms in ↑WS compared to ↑WS depression. A recent GWAS demonstrated that alcohol consumption and alcohol use disorders (AUD) show significant genetic differences, with the genetics of AUD being more closely related to other psychiatric disorders, and the genetics of alcohol consumption to that of some positive health outcomes, such as reduced risk of cardiovascular disease, and lower BMI, in line with our findings (Kranzler et al., 2019).

Taken together, these results suggest partially distinct genetic pathways between depression with atypical and typical neurovegetative symptoms and that this divergence may be attributable to distinct genetic predisposition to immune-metabolic traits. Although there is no convincing evidence that these depression subgroups may respond differently to conventional antidepressant treatments, drugs acting on the specific biological mechanisms implicated in ↑WS depression may have clinical benefits. For example, peroxisome proliferator-activated receptor (PPAR)-γ agonists target insulin resistance and the related oxidative and pro-inflammatory changes (which are also involved in the pathogenesis of depressive symptoms); they were demonstrated to have antidepressant effects in patients with treatment-resistant resistant bipolar depression and concomitant insulin resistance (Kemp et al., 2014).

The SNP-based heritability was estimated to be similar between ↑WS depression and other subtypes and it did not significantly change after excluding cases with probable bipolar disorder and schizophrenia. GCTA-GREML 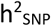 was similar to a previous study of depression with weight/appetite gain 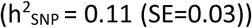 and weight/appetite loss 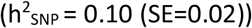 (z=1.24, p=0.22; z=1.72, p=0.09, respectively) (Milaneschi et al., 2017). However, this study considered both weight and appetite changes in the definition of the subgroups, while we used only weight changes because of the lack of information on the direction of appetite changes in UKB. Genetic correlations suggested that ↑WS depression genetics is highly shared with depression without ↑WS or ↑WS, and the genetic correlation between these subgroups was significantly higher than the genetic correlation between ↑WS depression and ↓WS depression (z=2.49, p=0.013). In line with previous evidence, major depression subgroups did not show evidence of negative natural selection based on GCTB-Bayes S estimates (Zeng et al., 2018).

Observational studies have shown differential pathophysiological correlates among subtypes of depression, with atypical depression more strongly associated with obesity and metabolic dysregulations than other depression groups (Brailean et al., 2020) (Lasserre et al., 2014) (Milaneschi et al., 2017). Consistent with previous research, the present findings suggest that the phenotypic association between depression with atypical features and cardiometabolic traits may partly result from shared genetic and biological mechanisms (Milaneschi et al., 2016) (Milaneschi et al., 2017). For example, pleiotropy may occur when shared genetic variants influence obesity-related traits and ↑WS depression through common pathways such as inflammation and/or leptin system dysregulation (Guo & Lu, 2014) (Ring & Zeltser, 2010). A previous study (Milaneschi et al., 2017) reported that the genetic correlation between leptin and ↑WS depression was decreased but not absent when considering leptin adjusted for BMI, while we found no association between PRS for BMI-adjusted leptin levels and ↑WS depression or depression with weight increase. Leptin reduces food intake and obesity is associated with increased leptin levels, through the induction of leptin-resistance (Myers et al., 2010). The adjustment of leptin levels for BMI eliminates an important source in leptin inter-individual variability, and separates the genetic factors regulating BMI from those that regulate only leptin production. Our results indicate that the genetic factors predisposing to increased BMI and increased leptin levels in ↑WS depression are highly correlated, in line with the hypothesis that increased leptin levels are generally a consequence of increased BMI rather than a cause (at least in common forms of obesity), although different pathogenetic mechanisms may lead to similar phenotypes (Myers et al., 2010). This discrepancy may also relate to the different criteria used to define depression subgroups and sample-specific characteristics.

The current findings also showed that PRS for alcohol daily use was inversely associated with risk for ↑WS depression both in case only and case-control analyses, while it showed no difference between ↓WS depression and healthy controls. In case-control comparisons, the PRS of alcohol dependence was associated with ↓WS depression and at the nominal level with ↑WS depression. Epidemiological studies have shown increased alcohol consumption and/or risk of alcohol use disorders among persons with atypical depression (Blanco et al., 2012) (Brailean et al., 2020). There are no previous studies which examined the genetic overlap between alcohol use and atypical depression, although a positive genetic correlation between MDD and alcohol dependence has been reported (Andersen et al., 2017) (Kranzler et al., 2019). Our results suggest that genetic risk for daily alcohol use is lower in ↑WS depression vs other groups, in contrast with epidemiological observations of higher risk of alcohol use disorders in atypical cases. However, as previously noted, the genetics of alcohol daily use only partially overlaps with the genetics of alcohol dependence. In addition, the increased rate of alcohol use among cases with ↑WS depression may result from secondary or environmental risk factors rather than from a genetic aetiology. In line with this hypothesis, depression with atypical features in UKB show longer, more severe and recurrent disease episodes, increased risk of comorbidities, and more lifetime deprivation and adversity (Brailean et al., 2020), and these factors may be responsible for increased risk of alcohol use disorders (Yang et al., 2018).

Several limitations should be considered when interpreting the results from the present study. First, the UK Biobank is not representative of the general population, with respondents more likely to be older, female, healthier, of a higher socioeconomic background and better educated (Fry et al., 2017). Compared to the overall UKB cohort, respondents of the MHQ have on average higher educational and occupational attainment, are less likely to smoke, or report longstanding illness/disability (Davis & Hotopf, 2019). This is likely to impact on prevalence and severity of depression within the sample, with persons with severe depression less likely to have completed the MHQ. Second, measures for depression were based on recall of the worst episode of depression and assessed using self-reported symptoms, rather than clinical diagnoses. This raises the possibility that self-reported symptoms may be affected by recall bias or other medical conditions, and we are not able to exclude that participants had other depressive episodes with different neurovegetative symptoms compared to the one reported in the MHQ. Third, our measurement of depression with atypical features did not capture all DSM-5 symptoms but only the neurovegetative component. Our results were in line with previous studies and contributed to characterise the genetics of these symptoms, but future research using the full atypical spectrum should be considered in future research and dimensional classifications should be considered as an alternative to binary categorisation. Fourth, due to the small numbers of individuals of non-European groups in UK biobank, and their systematic under-representation in GWAS studies used to derive PRS, our sample is limited to individuals of European descent, limiting the generalisability of our findings. Last, we had no statistical power to perform a genome-wide association study of ↑WS depression or Mendelian randomization to assess the causal relationship between immune-metabolic traits and ↑WS depression.

In conclusion, this study showed specific genetic overlap of ↑WS and ↑WS depression with immune-metabolic and alcohol-related traits. Our findings suggest that depression with typical and atypical neurovegetative symptoms may represent relatively homogenous subtypes characterised by partially distinct genetic liabilities. Understanding the shared genetic aetiology between immune-metabolic traits and depression subtypes could play an important role in the prevention/treatment of depressive episodes and the development of tailored treatments.

### Financial support

This research was supported by the UK Medical Research Council (MR/N015746 and MR/S0151132). This paper represents independent research part-funded by the National Institute for Health Research (NIHR) Biomedical Research Centre at South London and Maudsley NHS Foundation Trust and King’s College London. The views expressed are those of the author(s) and not necessarily those of the NHS, the NIHR or the Department of Health and Social Care.

Chiara Fabbri is supported by Fondazione Umberto Veronesi (https://www.fondazioneveronesi.it).

## Data Availability

UK Biobank data was accessed under application 18177.

## Acknowledgements

The authors acknowledge use of the research computing facility at King’s College London, Rosalind (https://rosalind.kcl.ac.uk), which is delivered in partnership with the National Institute for Health Research (NIHR) Biomedical Research Centres at South London & Maudsley and Guy’s & St. Thomas’ NHS Foundation Trusts, and part-funded by capital equipment grants from the Maudsley Charity (award 980) and Guy’s & St. Thomas’ Charity (TR130505).

UK Biobank data was accessed under application 18177 “Multi-trait GWAS analyses in the UK Biobank”.

We thank the CHARGE Inflammation Working Group (CIWG), the Psychiatric Genomics Consortium (PGC), the International Cannabis Consortium (ICC), the Genetics of Personality Consortium (GPC), the DIAbetes Genetics Replication And Meta-analysis (DIAGRAM) Consortium, the Global Lipids Genetics Consortium (GLGC), The ADIPOGen Consortium, The AGEN-BMI Working Group, The CARDIOGRAMplusC4D Consortium, The CKDGen Consortium, The ICBP, The MAGIC Investigators, The MuTHER Consortium, The MIGen Consortium, The PAGE Consortium, The ReproGen Consortium, The GENIE Consortium, The International Endogene Consortium, AFGen Consortium, Cohorts for Heart and Aging Research in Genomic Epidemiology (CHARGE) Consortium, International Genomics of Blood Pressure (iGEN-BP) Consortium, INVENT Consortium, STARNET, COMPASS Consortium, EPIC-CVD Consortium, EPIC-InterAct Consortium, International Stroke Genetics Consortium (ISGC), METASTROKE Consortium, Neurology Working Group of the CHARGE Consortium, NINDS Stroke Genetics Network (SiGN), UK Young Lacunar DNA Study, MEGASTROKE Consortium and all the authors that contributed to generate the summary statistics used for the analyses of this study.

